# A survey of patient and public perceptions and awareness of SARS-CoV-2-related risks among participants in India and South Africa

**DOI:** 10.1101/2022.08.26.22279242

**Authors:** Oluchi Mbamalu, Surya Surendran, Vrinda Nampoothiri, Candice Bonaconsa, Fabia Edathadathil, Nina Zhu, Vanessa Carter, Helen Lambert, Carolyn Tarrant, Raheelah Ahmad, Adrian Brink, Ebrahim Steenkamp, Alison Holmes, Sanjeev Singh, Esmita Charani, Marc Mendelson

## Abstract

A cross-sectional survey was performed among the adult population of participating countries, India and South Africa. The purpose of this study was to explore perceptions and awareness of SARS-CoV-2-related risks in the relevant countries. The main outcome measures were the proportion of participants aware of SARS-CoV-2, and their perception of infection risks.

Self-administered questionnaires were used to collect data via a web- and paper-based survey over three months. For data capturing, Microsoft Excel was employed, and descriptive statistics used for presenting data. Pearson’s Chi-squared test was used to assess relationships between variables, and a p-value less than 0.05 was considered significant.

There were 844 respondents (India: n=660, South Africa: n=184; response rate 87.6%), with a 61.1% vs 38.3% female to male ratio. Post-high-school or university education was the lowest qualification reported by most respondents in India (77.3%) and South Africa (79.3%). Sources of information about the pandemic were usually media and journal publications (73.2%), social media (64.6%), family and friends (47.7%) and government websites (46.2%). Most respondents correctly identified infection prevention measures (such as physical distancing, mask use), with 90.0% reporting improved hand hygiene practices since the pandemic. Hesitancy or refusal to accept the SARS-CoV-2 vaccine was reported among 17.9% and 50.9% of respondents in India and South Africa, respectively. Reasons cited included rushed vaccine development and the futility of vaccines for what respondents considered a self-limiting flu-like illness.

Respondents identified public health promotion measures for SARS-CoV-2. Reported hesitancy to the up-take of SARS-CoV-2 vaccines was much higher in South Africa. Vaccination campaigns should consider robust public engagement and contextually fit communication strategies with multimodal, participatory online and offline initiatives to address public concerns, specifically towards vaccines developed for this pandemic and general vaccine hesitancy.

## INTRODUCTION

The SARS-CoV-2 pandemic has highlighted the importance of infection prevention at individual and community levels. The World Health Organization (WHO) has indicated that for public health infection prevention measures to be successful, all members of society (communities and professional groups included) should be fully engaged [1]. These measures include but are not limited to physical distancing, masking, hand hygiene, avoiding poorly ventilated indoor spaces, and isolation/quarantine if infected or exposed. For efficient buy-in and contribution to these measures, individuals should understand the risks, mode of viral transmission, and consequences of infection. As such, the success of infection prevention measures depends on individual and community-level awareness and the adoption of infection prevention behaviours, which in turn depends on their perceptions and cognizance of risk.

While effective public engagement has been highlighted as key to gaining buy-in [2-4], additional research is needed to explore public awareness, perceptions and behaviours about SARS-CoV-2 and how these may influence adherence to public health measures, especially in low- and middle-income countries (LMIC). India (lower-middle-income) and South Africa (upper-middle-income) [5] are countries with emerging economies where the SARS-CoV-2 pandemic has had a significant impact [6]. Redeploying the capacity within an existing research collaboration across participating sites in these two countries [7, 8], we investigated the public’s perceptions and awareness of SARS-CoV-2-related risks and infection prevention practices through analysis of data contributed by participants across the two countries.

## METHODS

### Study design

We conducted a cross-sectional web- and paper-based survey. Data were collected using a self-administered questionnaire. Any adult member (over 18 years old) of the public, who provided informed consent before participation, was eligible to participate.

### Study development

The study development followed the STROBE cross-sectional reporting guidelines [9], as shown in Table 1 under Supporting Information.

**Table 1.** Self-reported respondent demographics

| Characteristic | India | South Africa | Total |
| --- | --- | --- | --- |
|  | n (%) | n (%) | n (%) |
| Country of residence | 660 (78.2) | 184 (21.8) | 844 (100) |
| **Gender** | **(n=660)** | **(n=184)** | **(n=844)** |
| Male | 285 (43.2) | 38 (20.7) | 323 (38.3) |
| Female | 369 (55.9) | 146 (79.3) | 515 (61.1) |
| Prefer not to say | 5 (0.8) | 0 | 5 (0.6) |
| Missing | 1 (0.2) | 0 | 0 |
| **Age** | **n = 657 (%)** | **n = 184 (%)** | **n = 841 (%)** |
| Younger than 20 years | 41 (6.2) | 7 (3.8) | 48 (5.7) |
| 20 to 29 years | 657 (47.2) | 14 (7.6) | 324 (38.5) |
| 30 to 39 years | 133 (20.2) | 21 (11.4) | 154 (18.3) |
| 40 to 49 years | 82 (12.5) | 57 (31.0) | 139 (16.5) |
| 50 to 59 years | 49 (7.5) | 49 (26.6) | 98 (11.7) |
| 60 to 69 years | 30 (4.6) | 27 (14.7) | 57 (6.8) |
| 70 years and older | 12 (1.8) | 9 (4.9) | 21 (2.5) |
| **Regular water supply** | **n = 650 (%)** | **n = 176 (%)** | **n = 826 (%)** |
| Yes | 560 (86.2) | 172 (97.7) | 732 (88.6) |
| No | 90 (13.8) | 4 (2.3) | 94 (11.4) |
| **Education** | **n = 653 (%)** | **n = 175 (%)** | **n = 828 (%)** |
| Primary schooling | 28 (4.3) | 1 (0.5) | 29 (3.5) |
| Secondary schooling | 113 (17.3) | 25 (13.6) | 138 (16.7) |
| Post-high school | 291 (44.6) | 96 (52.2) | 387 (46.7) |
| Post-graduate degree | 219 (33.5) | 50 (27.2) | 269 (32.5) |
| Other | 2 (0.3) | 3 (1.6) | 5 (0.6) |
| **Employment** | **n = 633 (%)** | **n = 166 (%)** | **n = 799 (%)** |
| Student | 135 (21.3) | 5 (3.0) | 140 (17.5) |
| Employed, part time | 35 (5.5) | 11 (6.6) | 46 (5.8) |
| Employed, full time | 249 (39.3) | 62 (37.3) | 311 (38.9) |
| Self-employed | 47 (7.4) | 50 (30.1) | 97 (12.1) |
| Retired/Pensioner | 27 (4.3) | 19 (11.4) | 46 (5.8) |
| I was furloughed/laid off during the lockdown | 8 (1.3) | 5 (3.0) | 13 (1.6) |
| Unemployed | 122 (19.3) | 13 (7.8) | 135 (16.9) |
| Other | 10 (1.6) | 1 (0.6) | 11 (1.4) |

**Table 1 (Supporting Information):** Reporting Checklist for cross-sectional study

The research team – made up of pharmacists, physicians, nurses, social scientists, patient advocate and public engagement specialist, and quantitative data analysts – designed a 42-question survey to elicit information on the public’s knowledge, perceptions and awareness of SARS-CoV-2 infection risks. The 4-part survey included participant demographics, general knowledge of SARS-CoV-2, perceived risks and barriers, and self-efficacy. In South Africa, survey questions and participant information leaflets were translated into IsiZulu, IsiXhosa and Afrikaans languages, whereas in India, the paper-based survey was translated into Malayalam for local distribution. The survey was piloted with members of the public, and relevant revisions were made before dissemination.

### Study settings and participant recruitment

The survey was open for participation by any member of the public over a 3-month duration. Voluntary response sampling was utilized. All invited individuals received participant information leaflets, and those willing to participate had to provide informed consent before commencing the survey. Participation was voluntary across both countries.

In South Africa, the survey was available online in three languages – IsiXhosa, Afrikaans and English. In India, the survey was available online in the English language, and in the paper format in two languages, English and Malayalam.

### Data collection

Data collection took place from 15 September to 15 December 2020 and coincided with the first wave of the SARS-CoV-2 pandemic in India and the beginning of the second wave in South Africa. In South Africa, the survey was available online *via* Qualtric. In India, in addition to the online platform in English, paper surveys (in English and Malayalam) were also distributed among participants (patients, patient carers and/or visitors) at the study site (hospital) in Kerala.

### Ethics Statement

The study was approved by the relevant human research ethics committees at the Amrita Institute of Health Sciences, Kerala, India (Ref: IRB-AIMS-2020-232) and the University of Cape Town, South Africa (Ref: 311/2020). Formal consent was obtained prior to participation in the survey. For the online and paper versions of the survey, consent was indicated by the participant ticking the relevant box for consent on the survey form.

### Statistical analysis

Data from participants who completed the paper-based format were captured in a Microsoft (MS) Excel file and codes assigned, while data of participants who completed the online form were exported to MS Excel. The data from the paper-based and online versions of the survey were cleaned and combined.

Descriptive statistics were used to report participant characteristics and survey responses. The underlying outcomes were awareness of the pandemic, perceived threats and barriers, and self-efficacy. Responses were captured as categorical variables, reported as percentages of received feedback for each item of interest (missing data were excluded) or, where possible, data were scaled from strongly agree to strongly disagree. Pearson’s Chi-squared test was used to assess relationships between variables, and p < 0.05 was considered statistically significant.

## RESULTS

### Participant demographics

There was a total of 844 respondents (660 participants from India and 184 participants from South Africa). There were 318 respondents to the online survey and 342 patients or patient carer respondents to the paper survey in India. The response rate for the online survey was 87.6% (502/573), calculated as the ratio of participants who clicked on the survey link versus those who commenced participation. The response rate for the paper version of the survey could not be estimated, as respondents returned a higher number of the completed survey forms than the initial number disseminated, indicating the forms had been copied and shared more widely.

There were more female (515/844, 61.0%) than male (323/844, 38.3%) respondents (Table 1). Three entries for age were excluded (one was invalid with two selections and two were missing), resulting in a total response of 657 for age entries. Most of the respondents in India and South Africa were in the 20-29-year (310; [n=657] 47.2%) and 40-49-year (57; 31.0%) age groups, respectively.

The percentage of student respondents was higher in India (21.3%, 135/633) than South Africa (3.0%, 5/166). Unemployment was higher among respondents in India (19.3%, 122/163) than in South Africa (7.8%, 13/166), while there were more self-employed (30.1%, 50/166) and retired (11.4%, 19/166) respondents in South Africa.

### Knowledge and concerns of SARS-CoV-2 transmission and infection

Reported sources of SARS-CoV-2 information, completed by 652 and 172 participants in India and South Africa, respectively, are shown in Figures 1a and 1b. Media and journal publications were the most common sources of information, along with social media, family and friends, and government websites. On social media across both countries, Facebook®, WhatsApp® and YouTube® were the most frequently used sites for information about the pandemic.

**Figure 1a.**
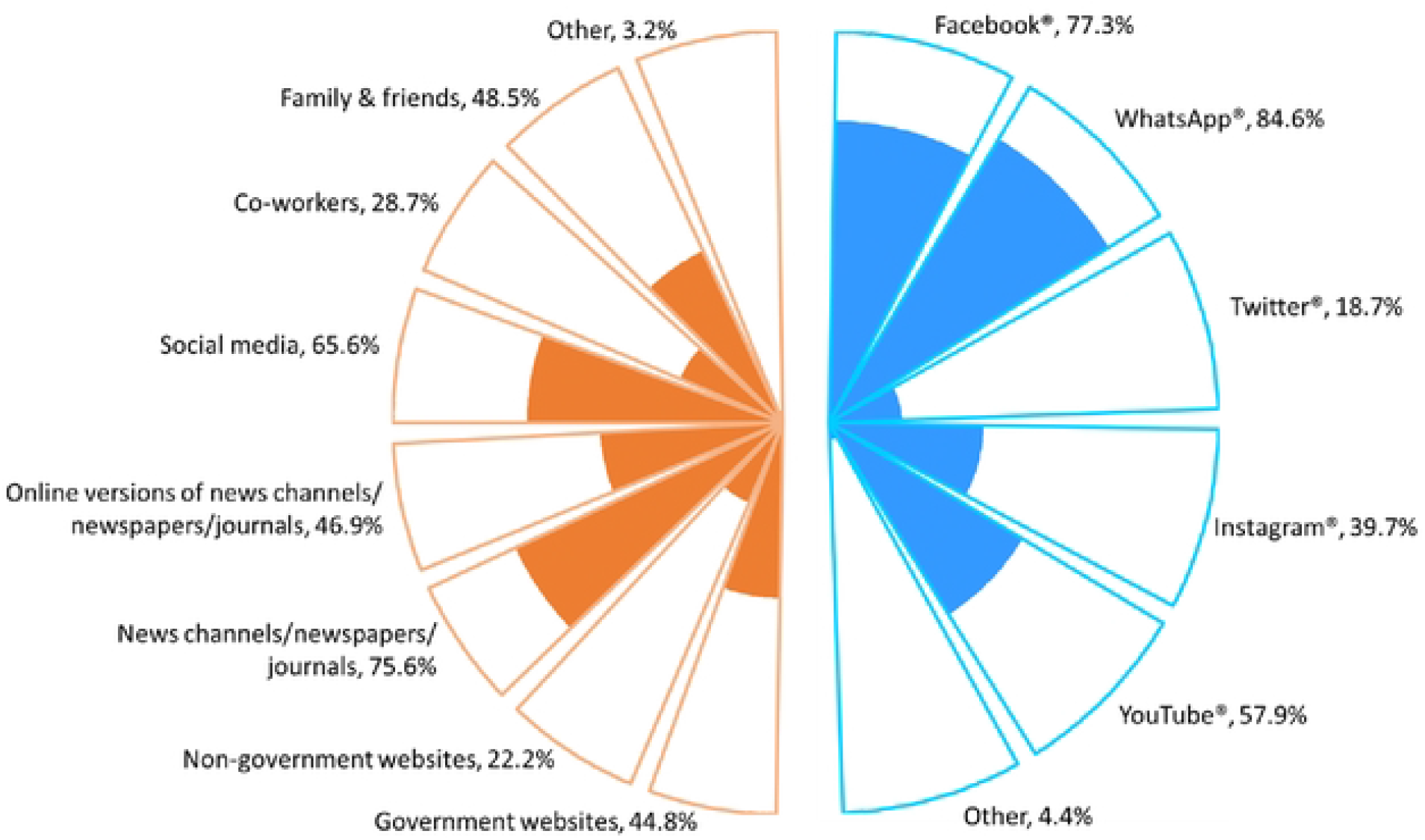
Respondents’ sources of SARS-CoV-2 information (traditional and social media) in India (n=652)

**Figure 1b.**
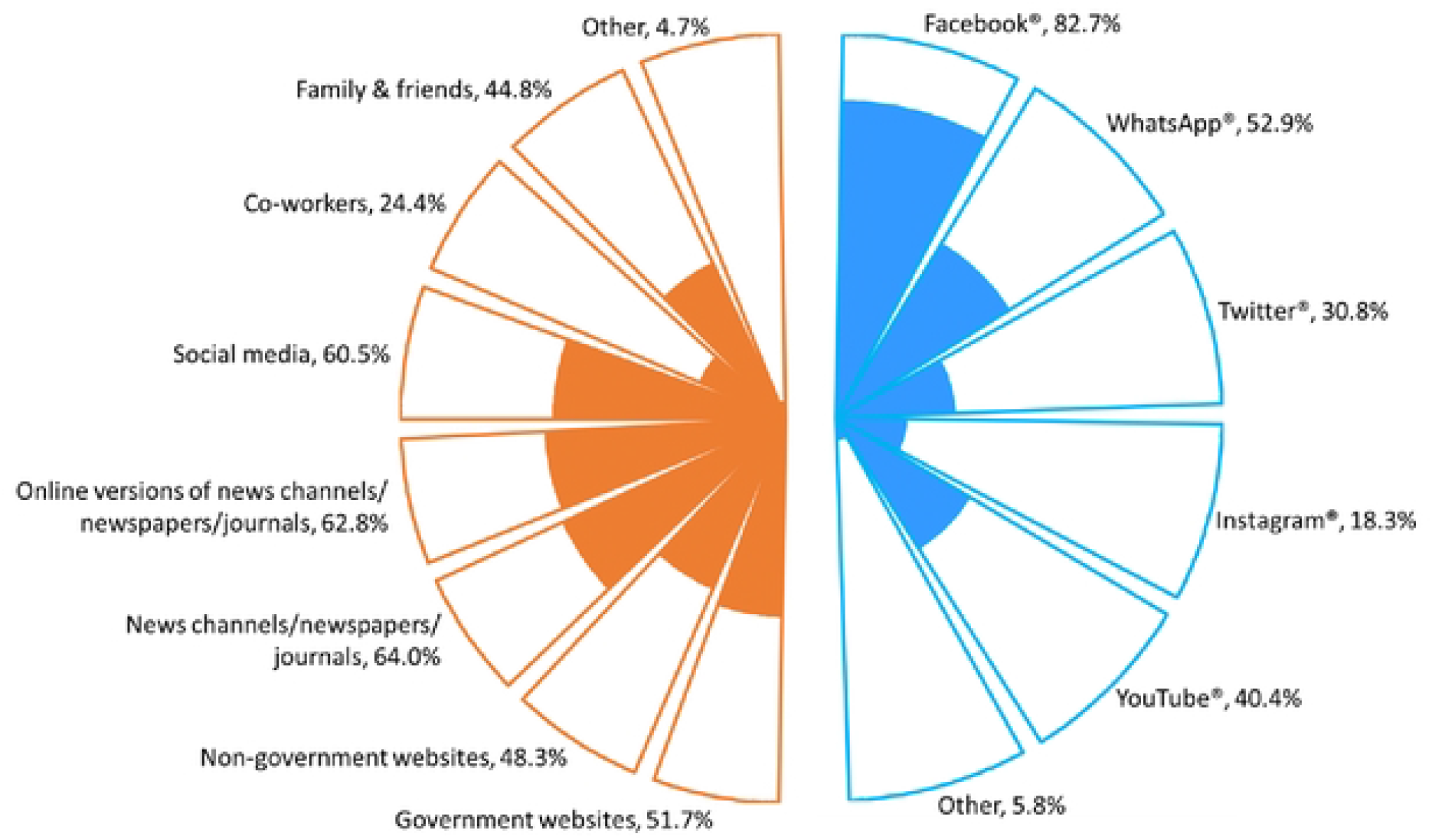
Respondents’ sources of SARS-CoV-2 information (traditional and social media) in South Africa (n=172)

In Table 2, the respondents’ knowledge of SARS-CoV-2 transmission routes, infection course and prevention/management options is summarised. The primary route of SARS-CoV-2 transmission identified was nasal/oral droplets, airborne particles, and infected body fluids. More than half of the respondents also demonstrated knowledge of SARS-CoV-2 incubation and symptom manifestation, quarantine objectives, and general duration of isolation for infected patients.

**Table 2.** Respondent’s knowledge and experiences of the pandemic

| Knowledge of SARS-CoV-2 | India | Response (%)<br>South Africa | Total |
| --- | --- | --- | --- |
| **Major routes of transmission** |  |  |  |
| Infected bodily fluids | 426/516 (82.6) | 55/119 (46.2) | 481/635 (75.7) |
| Nasal or oral droplets | 555/591 (93.9) | 158/165 (95.8) | 713/756 (94.3) |
| Airborne | 352/458 (76.9) | 114/147 (77.6) | 466/605 (77.0) |
| Foodborne | 113/328 (34.5) | 9/101 (8.9) | 122/429 (28.4) |
| Waterborne | 119/322 (37.0) | 5/98 (5.1) | 124/420 (29.5) |
| Other (please specify) | 11/60 (18.3) | 5/ 25 (20.0) | 16/85 (18.8) |
| **Time to symptom onset** | n = 626 (%) | n = 171 (%) | n = 797 (%) |
| Immediately – there is no delay | 42 (6.7) | 1 (0.6) | 43 (5.4) |
| 0 to 2 weeks | 448 (71.6) | 152 (88.9) | 600 (75.3) |
| 2 to 4 weeks | 80 (12.8) | 12 (7.0) | 92 (11.5) |
| Over 4 weeks | 10 (1.6) | 2 (1.2) | 12 (1.5) |
| I don't know | 35 (5.6) | 4 (2.3) | 39 (4.9) |
| Multiple entries | 11 (1.8) | 0 | 11 (1.4) |
| **Perceived reason for quarantine of SARS-CoV-2-positive individuals** | n = 625 (%) | n = 171 (%) | n = 796 (%) |
| To help them get better | 22 (3.5) | 0 | 22 (2.8) |
| To prevent them from infecting others | 430 (68.8) | 124 (72.5) | 554 (69.6) |
| There is no good reason for that | 3 (0.5) | 12 (7.0) | 15 (1.9) |
| Other, please specify | 1 (0.2) | 2 (1.2) | 3 (0.4) |
| I don't know | 9 (1.4) | 0 | 9 (1.1) |
| Multiple entries | 159 (25.4) | 33 (19.3) | 192 (24.1) |
| **Duration of isolation (if not admitted to a healthcare facility)** | n = 625 (%) | n = 171 (%) | n = 796 (%) |
| As soon as coughing stops | 5 (0.8) | 0 | 5 (0.6) |
| 10 to 14 days after symptoms first started | 314 (50.0) | 136 (79.5) | 450 (56.5) |
| As soon as they feel better | 36 (5.7) | 3 (1.8) | 39 (4.9) |
| 21 days after symptoms stop | 110 (17.5) | 9 (5.3) | 119 (14.9) |
| For asymptomatic cases: as advised by healthcare guidelines | 61 (9.7) | 10 (5.8) | 71 (8.9) |
| They do not need to be isolated | 3 (0.5) | 9 (5.3) | 12 (1.5) |
| I don't know | 28 (4.5) | 4 (2.3) | 32 (4.0) |
| Multiple entries | 71 (11.3) | 0 | 71 (8.9) |
| **Changes in hand washing practices** | n = 615 (%) | n = 167 (%) | n = 782 (%) |
| I wash/sanitise my hands more often | 561 (91.2) | 143 (85.6) | 704 (90.0) |
| I wash/sanitise my hands less often | 13 (2.1) | 0 | 13 (1.7) |
| There is no difference in how often I wash/sanitise my hands | 18 (2.9) | 23 (13.8) | 41 (5.2) |
| Other, please specify | 2 (0.3) | 1 (0.6) | 3 (0.4) |
| I don't know | 6 (1.0) | 0 | 6 (0.8) |
| Multiple entries | 15 (2.4) | 0 | 15 (1.9) |
| **Avoided visit to healthcare facility because of SARS-CoV-2-** | n = 594 (%) | n = 161 (%) | n = 755 (%) |
| Yes | 171 (28.8) | 60 (37.3) | 231 (30.6) |
| No | 353 (59.4) | 70 (43.5) | 423 (56.0) |
| Not applicable/had no need to visit a healthcare facility | 70 (11.8) | 31 (19.3) | 101 (13.4) |
| Would you have a SARS-CoV-2 vaccination? | n = 587 (%) | n = 161 (%) | n = 748 (%) |
| Yes | 482 (82.1) | 79 (49.1) | 561 (75.0) |
| No | 36 (6.1) | 55 (34.2) | 91 (12.2) |
| I don't know | 69 (11.8) | 27 (16.8) | 96 (12.8) |

More frequent hand washing was reported across both countries (90.0%); however, a higher percentage of respondents in South Africa (13.8%) than in India (2.9%) noted no difference in their hand hygiene practices. Overall, 75.0% of all the respondents indicated their willingness to receive vaccination when it becomes available; however, the percentages were higher in India (82.1%) than in South Africa (49.1%). The most common reasons cited for apathy to vaccination were perceptions of rushed vaccine development and the futility of vaccines for what respondents considered a self-limiting flu-like illness.

### Self-efficacy: perceptions of SARS-CoV-2 infection prevention measures

Respondents’ perceptions and concerns about their ability to cope with SARS-CoV-2 infection prevention measures are presented in Figure 2, given their perceived knowledge and awareness of the pandemic and infection risks. More than half of respondents in each country reported that they have sufficient knowledge of SARS-CoV-2, understood available information on the pandemic, would know what to do or questions to ask if they or someone else contracted SARS-CoV-2, and have access to healthcare were they to become ill with SARS-CoV-2 infection, and would be able to cope with extended containment measures such as a lockdown. Compared to South Africa, more respondents in India reported concern over infection, its financial implications and associated stigma. On the intent to wear a face mask, 8.6% and 26.2% of respondents in India and South Africa reported dissatisfaction with this measure while outdoors, respectively.

**Figure 2:**
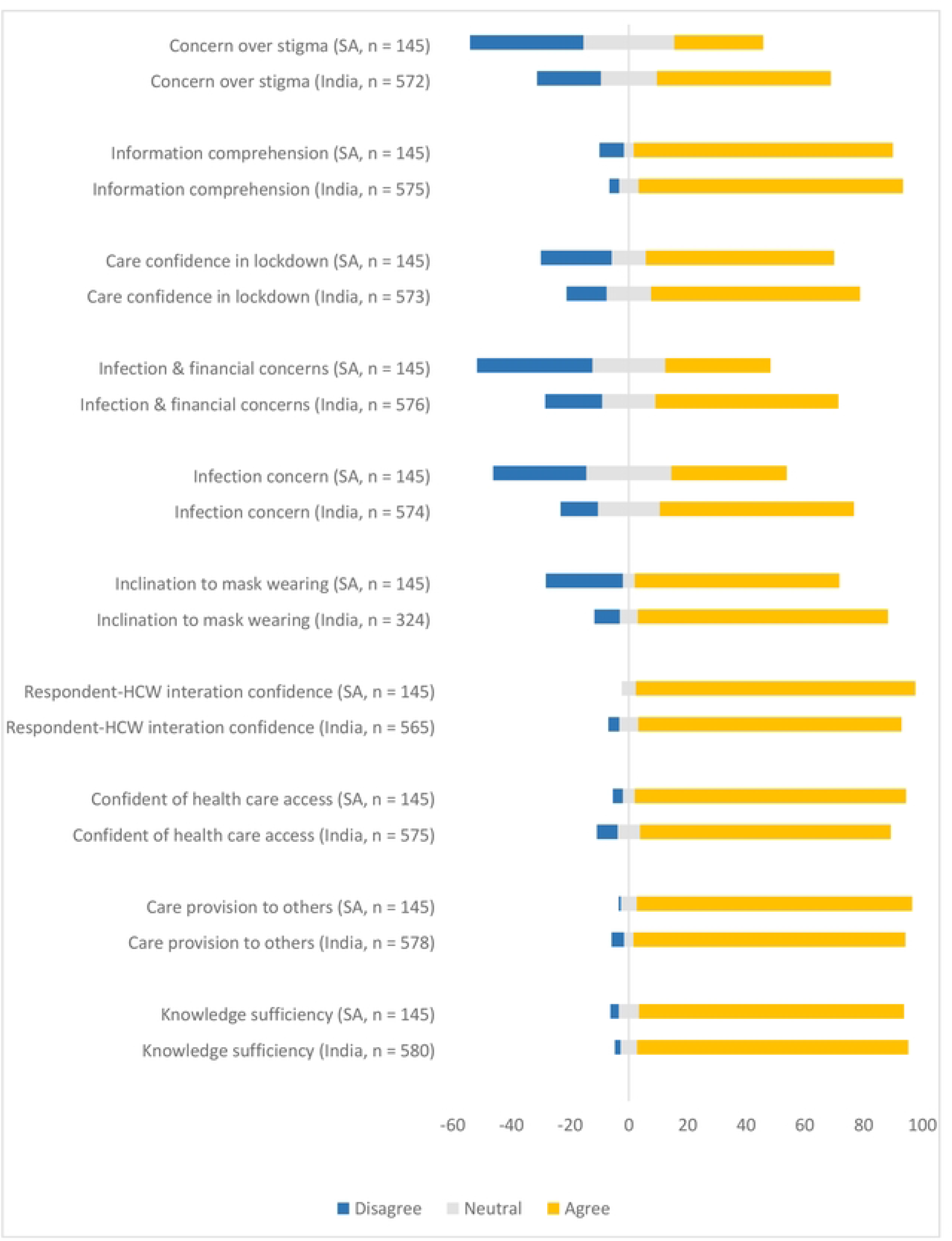
Respondents’ perceptions of self-efficacy in relation to coping with the COVID-19 pandemic in South Africa (SA) and India

There was no statistical significance between hand washing and water supply (Table 3), as even those without access to water supply reported that they washed their hands more frequently since the pandemic (p = 0.2168 and p = 0.7970 in India and South Africa, respectively). Water supply showed a mixed relationship with employment as some full-time workers had no access to water. The test highlights a difference between participants in the two countries; p = 0.008 and 0.4471 for India and South Africa, respectively.

**Table 3.** Relationships between selected variables

| A. Is hand washing affected by water supply? |  |  |  |  |
| --- | --- | --- | --- | --- |
| Hand wash frequency | India |  | South Africa |  |
|  | Yes | No | Yes | No |
| Wash more | 476 | 79 | 140 | 3 |
| Wash less | 13 | 0 | 0 | 0 |
| No change | 15 | 2 | 22 | 1 |
| Other | 2 | 0 | 1 | 0 |
| Don't know | 5 | 1 | 0 | 0 |
| Multiple | 10 | 5 | 0 | 0 |
| p-value | 0.2168 |  | 0.797 |  |

| B. Is water supply affected by employment? |  |  |  |  |
| --- | --- | --- | --- | --- |
| Employment Status | India |  | South Africa |  |
|  | Yes | No | Yes | No |
| Student | 124 | 9 | 5 | 0 |
| Part time | 27 | 8 | 11 | 0 |
| Full time | 217 | 29 | 62 | 0 |
| Self-employed | 40 | 7 | 47 | 3 |
| Unemployed | 101 | 20 | 12 | 1 |
| Retired | 18 | 8 | 19 | 0 |
| Other | 8 | 2 | 1 | 0 |
| Laid off | 4 | 4 | 5 | 0 |
| *p-value* | *0.0008026* |  | *0.4471* |  |

| C. Is avoidance of healthcare facilities because of fear of COVID-19 contraction influenced by age? |  |  |  |  |  |  |
| --- | --- | --- | --- | --- | --- | --- |
| Age | India |  |  | South Africa |  |  |
|  | Yes | No | N/A | Yes | No | N/A |
| <20 | 11 | 15 | 11 | 2 | 1 | 1 |
| 20-29 | 83 | 147 | 42 | 4 | 4 | 2 |
| 30-39 | 28 | 81 | 9 | 7 | 9 | 4 |
| 40-49 | 24 | 49 | 4 | 21 | 18 | 12 |
| 50-59 | 11 | 31 | 3 | 17 | 19 | 8 |
| 60-69 | 10 | 19 | 1 | 8 | 14 | 3 |
| >=70 | 2 | 10 | 0 | 1 | 5 | 1 |
| *p-value* | *0.001451* |  |  | *0.9008* |  |  |

| D. Is avoidance of health care facilities because of fear of COVID-19 contraction influenced by employment? |  |  |  |  |  |  |
| --- | --- | --- | --- | --- | --- | --- |
| Employment Status | India |  |  | South Africa |  |  |
|  | Yes | No | N/A | Yes | No | N/A |
| Student | 37 | 57 | 27 | 2 | 1 | 1 |
| Part time | 12 | 20 | 1 | 5 | 3 | 3 |
| Full time | 57 | 137 | 27 | 25 | 21 | 11 |
| Self-employed | 11 | 28 | 2 | 10 | 26 | 10 |
| Unemployed | 31 | 79 | 5 | 5 | 6 | 1 |
| Retired | 9 | 17 | 0 | 8 | 7 | 2 |
| Other | 5 | 3 | 1 | 0 | 1 | 0 |
| Laid off | 1 | 4 | 3 | 0 | 2 | 2 |
| *p-value* | *0.0002084* |  |  | *0.4076* |  |  |

Our results show that some respondents avoided healthcare facilities during this pandemic. Some participants in this study, particularly in India, reported avoiding healthcare facilities because of a fear of contracting the COVID-19 virus; this was affected by employment status, more in India (p = 0.0002) than in South Africa (p = 0.4076).

## DISCUSSION

This study provides insight into the public’s awareness and perspectives of the SARS-CoV-2 infection and risks in two middle-income countries hard hit by the pandemic [6]. The aim was to gain some understanding of the knowledge and views about the pandemic, particularly when considering the expected roles that the public have in this pandemic regarding social distancing and infection prevention through hand hygiene, mask use and vaccination uptake.

At the time of the study, these two countries were at different phases of the pandemic infection curves with no viable vaccines available. Although these data are somewhat dated, these findings add to the body of knowledge on the public’s perceptions of the pandemic. Also, how a better understanding of this information can be leveraged for improved infection prevention and behavioural interventions and promotions for this and the future infectious disease pandemics. Such knowledge will be helpful in infectious disease pandemic control and mitigation, including in the ongoing COVID-19 pandemic. The insight from this study can assist with measures to address continued vaccine hesitancy and inequity when many countries are dealing with a fourth or subsequent infection wave.

From the onset of the pandemic, efforts have been communicated to inform the public of infection risks and required containment/mitigation measures. The need for public engagement and hygiene intervention, behaviour change, and consideration of socio-cultural aspects in public awareness initiatives in India and South Africa has been noted in the literature [10-14]. The volume of news media dedicated to the pandemic may also have served to provide education and awareness among the public. Conversely, it may have fuelled confusion and panic, particularly on the diverse online channels where unbridled and unverified evidence and opinions compete for attention with information from local and global health authorities.

Survey respondents demonstrated awareness of the pandemic, with most identifying the primary routes of transmission, incubation period, symptoms of infection, and recommended measures for infection prevention and management of mild conditions, including the reason for and duration of isolation. Information on the pandemic was generally gained from traditional and social media, family and friends, and government websites. Respondents’ reliance on general and social media as sources of pandemic-related information highlights the role played by the media in pandemic containment and mitigation.

The information landscape has changed extensively in the last three decades, prompting the need to address not only the SARS-CoV-2 pandemic but also its related infodemic [15]. While the main aspects of an infodemic refer to inaccurate and misleading information shared through digital and physical environments during disease outbreaks, disinformation refers to the deliberate spread of false information. In this pandemic, we are increasingly witnessing a growing infodemic driven by misinformation, including a worrying trend in the escalation of disinformation through traditional and social/digital media [16-18]. The role of the media, traditional and digital alike, in framing and rapidly disseminating information is evident in this pandemic, particularly when related to influencing behaviours and empowering individuals with the accurate information to make informed decisions regarding IPC [15, 19-21].

Family and friends were noted as sources of SARS-CoV-2 information by respondents in the survey. Word of mouth presented face-to-face or through various communication channels within families and among friends, though not specifically a media source, is an essential source of information. It is also a key route for spreading misinformation, mainly because of the trust between the source and the recipient. Thus, the prominence of influencers (in the community and on digital platforms alike) in disseminating pandemic-related information is highlighted.

The findings of this study underscore the importance of various media as sources of information for informed decision-making among the public. It also draws attention to the relevance of social media, and family and friends, as sources of pandemic-related information for the public. Given the infodemic that has trailed the SARS-CoV-2 pandemic on all media [21, 22], there is a need for evidence-informed and timely communication in continually addressing pandemic-related misinformation and disinformation. Infodemic management is multifaceted, requiring different disciplines to address it. Beyond communication, factors influencing an individual’s behaviours may relate to external pressures, including the economy, politics, education, health literacy and religious or cultural beliefs.

Some respondents in this study considered SARS-CoV-2 to be food- or water-borne. Such beliefs may impact infection prevention measures; while there has been research into transmission by these routes [23, 24], they have not been noted as primary transmission routes for the viral infection. Droplet and airborne transmission have been noted as some primary transmission routes, with the use of face masks a significant intervention in reducing the spread of the infection [25, 26].

Across both countries, some respondents expressed somewhat reluctance to mask-wearing, despite their concern about contracting the infection, which may be related to the stigma or discomfort of masks. Stigma, known to influence/compromise infection prevention behaviours [27, 28], needs to be addressed, locally and globally, not only for the current pandemic but also for future ones, and improved adherence to optimised infection prevention practices.

Among other options to reduce infection risk, hand hygiene has been prioritised in public health messages for pandemic mitigation [29]. Access to clean water is critical for hand hygiene and is among the tools to address and mitigate the impact of the pandemic, as highlighted in the literature [29, 30]. While water supply did not affect hand hygiene frequency among respondents, it highlighted a difference between study participants in the two countries. Infection prevention measures such as hand hygiene and physical distancing may pose a challenge in some LMIC (India and South Africa are examples), especially in under-resourced sections of rural areas or densely populated urban settings [29, 30].

Isolation and quarantine of infected and exposed individuals are underlying measures for infectious disease control, though this may prove challenging. Responses to SARS-CoV-2-related isolation/quarantine duration reflect respondents’ perceptions of SARS-CoV-2 incubation. While there was an initial consensus on a 14-day isolation/quarantine period for infected/affected individuals, there have been shifts and debates on the optimum incubation period of the virus, hence, the duration of isolation and quarantine measures [31]. Respondents’ responses reflected this, more so in India, where discussions about extended isolation periods have been reported [32].

Lockdown measures instituted in various parts of the world following the spread of SARS-CoV-2 served as another infectious disease mitigation strategy. With the rise of infection transmission and the attendant lockdown measures, it was expected that individuals would have avoided visiting healthcare facilities. Some participants in this study, particularly in India, reported avoiding healthcare facilities because of a fear of contracting the COVID-19 virus; this was influenced by employment status, more in India than in South Africa. Employed participants may be more likely motivated to maintain good health or hesitant to confirm illness, for fear of losing money or work, resulting in fewer visits to healthcare facilities, than those unemployed. While lockdown measures can reduce patient presentation to healthcare facilities [33, 34], such a decline in presentation may also be associated with later presentations with more severe consequences. Initiatives are required to address gaps in patient care necessitated by public health promotion strategies such as lockdowns in this and future pandemics.

Across the two countries, attitudes to the vaccination were positive. However, the country analysis showed this was driven by higher vaccine acceptance in India, with respondents in South Africa more cautious regarding COVID-19 vaccination. Reasons cited for hesitancy or a negative attitude to SARS-CoV-2 vaccination were related to mistrust in the vaccine development process and the futility of vaccines for what respondents considered a self-limiting flu-like illness.

This survey was, however, conducted before SARS-CoV-2 vaccines were available. Hesitancy towards the SARS-CoV-2 vaccine had been noted earlier in the pandemic, fuelled by infodemic on communication channels and the public’s belief in SARS-CoV-2-related conspiracies [35, 36]. The notion that the pandemic has been grossly exaggerated and reported, with unnecessary financial and other stresses on populations, was expressed by some participants who provided additional free text information. As the pandemic evolves, research to better understand infection and vaccine-related concerns among the general population is needed to support targeted and contextually appropriate strategies promoting vaccine uptake and optimised infection prevention behaviours.

Among individuals with opposing opinions about vaccination, using social science methods to study underlying reasons and contexts for their views, along with highlighting the individual rather than the collective advantages of vaccination, may provide helpful and relatable insight [36, 37]. This could be particularly important when considered in light of recent research and noted factors that may influence vaccine perception and uptake [38-40]. More recent research has provided insight into dealing with vaccine-hesitancy as well as the challenges associated with anti-vaxxers [41]. Public health campaigns and vaccination promotions should therefore understand and leverage social listening techniques to comprehend public perceptions concerning communication gaps. A similar method of social listening should be developed for community and traditional settings to understand why various beliefs and behaviours related to COVID-19 emerged.

## STRENGTHS AND LIMITATIONS

Our study provides unique insights into the public’s attitudes and practices across two LMIC during the early stages of this pandemic. The findings are subject to some limitations, which should be considered in its interpretation.

First, being a cross-sectional study, the relevance of the findings may change over time and with interventions, especially as subsequent waves of COVID-19 have been reported. Second, the online distribution of the survey and the limited paper version may have limited its reach, particularly under-representing individuals from diverse socio-economic levels. Third, data collection across both sites did not rely on the same methods, given the COVID-19 restrictions at the time of data collection, which likely influenced the sample sizes across the sites. Sample size may also have been influenced by survey fatigue, challenges with Internet access in LMIC, and other limitations associated with accessing and participating in the survey at the time of the survey roll-out.

Survey respondents are therefore not representative of the public in either of the two countries, limiting the generalizability of findings. In addition, this survey was conducted between September and December 2020, when both countries were at different phases of the SARS-CoV-2 pandemic. The differences in experiences across the countries may have influenced the responses provided.

Nevertheless, this paper fills a gap in the knowledge, awareness and attitudes of the public in India and South Africa towards IPC practices in the context of COVID-19 within the first year of the pandemic. It will be beneficial for charting public understanding and perception of the COVID-19 pandemic and provide informative data that can be employed for public engagement in other infectious disease control and mitigation across both sites and similar contexts. While this research presents the data for each country separately, it is not its intention to make any statistical comparisons between participants in the two countries. Despite that, the individual test on how employment affects water supply and the avoidance of healthcare settings during lockdown provided some insight on differences between participants in the two countries. Thus, the need for pandemic mitigation efforts to consider differences in context and subjects for the delivery of context-specific and appropriate interventions is highlighted.

## CONCLUSIONS

This study presents socio-economic and demographic data, which may influence public awareness and behaviour and further be explored in pandemic mitigation initiatives among the public in both countries. Survey respondents correctly identified public health promotion measures for SARS-CoV-2. Reported disinclination to mask-wearing and reported hesitancy for the uptake of SARS-CoV-2 vaccination highlight gaps that can be addressed for improved pandemic mitigation efforts. Further research to explore the outlook towards mask use and vaccination across both countries can provide more insight on factors influencing infection prevention and vaccine apathy. Vaccination campaigns should consider robust public engagement and more targeted communication strategies using tactics like social listening, with multimodal, participatory online and offline initiatives to address the infodemic that drives public concerns. Furthermore, this can contribute to developing a better understanding of the driving force behind vaccine hesitancy among different populations.

## Data Availability

Data is available on a secure online portal, and can be accessed on request or included as supplementary information, if preferred.

## Acknowledgements

The authors express appreciation to all survey participants and members of the public who participated in the review of or provided feedback on the survey tool, and to Ms Jean Fourie for review and editing of the manuscript.

## Author contributions

OM conceptualised and wrote the initial protocol for the study, with additional input and revision from CB, VC, HL, RA, SSingh and overall oversight by EC and MM. OM, SSurendran and EC coordinated the data collection with input from NZ, SSurendran, VN, and FE contributed to data capturing; OM, FE and ES contributed to the data analysis. OM wrote the first draft of the manuscript, with input from SSurendran and oversight from EC and MM. All authors contributed to subsequent revisions and approval of the final draft.

## Declarations

### Ethics approval and consent to participate

The study was approved by the relevant human research ethics committees at the Amrita Institute of Health Sciences, Kerala, India (Ref: IRB-AIMS-2020-232) and the University of Cape Town, South Africa (Ref: 311/2020).

### Patient and public involvement

A patient advocate and public engagement specialist/civil society champion was involved in the design of the study material and also contributed as an author. Members of the public participated in the review of the survey tool and provided feedback for its modification. For the online and paper versions of the survey, consent was indicated by the participant ticking the relevant box for consent on the survey form.

## REFERENCES

1. World Health Organization. COVID-19 Strategy Up Date. 2020. Available: https://www.who.int/docs/default-source/coronaviruse/covid-strategy-update-14april2020.pdf [accessed 17 August 2020].

2. Woke FI. Communicating COVID-19 prevention health messages: a case study of South Africa. WebMed Central. Published online 2020:11(9): WMC005633. Available: https://www.webmedcentral.com/wmcpdf/Article_WMC005633.pdf [accessed 8 August 2021].

3. Gilmore B, Ndejjo R, Tchetchia A, de Claro V, Mago E, Diallo AA, et al. Community engagement for COVID-19 prevention and control: a rapid evidence synthesis. BMJ Glob Health 2020;5(10):1–11. doi:10.1136/bmjgh-2020-003188

4. Abdullahi L, Onyango JJ, Mukiira C, Wamicwe J, Githiomi R, Kariuki D, et al. Community interventions in low—and middle-income countries to inform COVID-19 control implementation decisions in Kenya: a rapid systematic review. PLoS One. 2020;15(12 December):1–29. doi:10.1371/journal.pone.0242403

5. World Bank. World Bank Country and Lending Groups – World Bank Data Help Desk. World Bank. Published online 2019:1–8. Available: https://datahelpdesk.worldbank.org/knowledgebase/articles/906519-world-bank-country-and-lending-groups [accessed 21 June 2021].

6. World Health Organization (WHO). COVID-19 Weekly Epidemiological Update 54. 2021 August 24. Available: https://www.who.int/publications/m/item/weekly-epidemiological-update-on-covid-19---24-august-2021 [accessed 29 August 2021].

7. Veepanattu P, Singh S, Mendelson M, Nampoothiri V, Edathadatil F, Surendran S, et al. Building resilient and responsive research collaborations to tackle antimicrobial resistance—lessons learnt from India, South Africa, and UK. Int J Infect Dis. 2020;100(P278-282). doi: https://doi.org/10.1016/j.ijid.2020.08.057

8. Singh S, Mendelson M, Surendran S, Bonaconsa C, Mbamalu O, Nampoothiri V, et al. Investigating infection management and antimicrobial stewardship in surgery: a qualitative study from India and South Africa. Clin Microbiol Infect. Published online 2021:1–27. doi:10.1016/j.cmi.2020.12.013

9. Von Elm E, Altman DG, Egger M, Pocock SJ, Gøtzsche PC, Vandenbroucke JP; STROBE Initiative. The Strengthening the Reporting of Observational Studies in Epidemiology (STROBE) statement: guidelines for reporting observational studies. J Clin Epidemiol. 2008;61(4):344–349. doi: 10.1016/j.jclinepi.2007.11.008

10. Bhatia R. Public engagement is key for containing COVID-19 pandemic. Indian J Med Res. 2020;151(2):118–120. Available: http://dx.doi.org/10.4103/ijmr.IJMR_780_20

11. Fedorowicz M, Arena O, Burrowes K. Community Engagement During The COVID-19 Pandemic And Beyond: A Guide For Community-Based Organizations. 2020 (September):1–33. Available: https://www.urban.org/research/publication/community-engagement-during-covid-19-pandemic-and-beyond/view/full_report [accessed 8 August 2020].

12. Department of Health (Republic of South Africa). An analysis of social behavioral change and the impact on social interactions. SAcoronavirus.co.za. Published online 2020a:1–12. Available: https://sacoronavirus.co.za/2020/08/20/an-analysis-of-social-behavioral-change-and-the-impact-on-social-interactions/

13. Malik A, Khan LM, Quan-Haase A. Public health agencies outreach through Instagram during COVID-19 pandemic: crisis and emergency risk communication perspective. Int J Disaster Risk Reduct. 2021;61, 102346. Available: https://ir.lib.uwo.ca/cgi/viewcontent.cgi?article=1368&context=fimspub

14. Department of Health (Republic of South Africa). COVID-19 national public hygiene strategy and implementation plan. 2020b. Available: ‘https://www.knowledgehub.org.za/system/files/elibdownloads/2020-04/covid-19%20national%20public%20hygiene%20strategy%20and%20implementation%20plan%281%29.pdf [accessed 8 August 2020].

15. Lep Ž, Babnik K, Hacin Beyazoglu K. Emotional responses and self-protective behavior within days of the COVID-19 outbreak: the promoting role of information credibility. Front Psychol. 2020;11(July):1–8. doi:10.3389/fpsyg.2020.01846

16. Parikh PA, Shah B V, Phatak AG, Vadnerkar AC, Uttekar S, Thacker N, et al. COVID-19 pandemic: knowledge and perceptions of the public and healthcare professionals. Cureus. 2020;12(5). doi:10.7759/cureus.8144

17. Wasserman H, Madrid-Morales D. Social Media Users In Kenya And South Africa Trust Science, But Still Share COVID-19 Hoaxes. The Conversation, 6 April 2021. Available: https://theconversation.com/social-media-users-in-kenya-and-south-africa-trust-science-but-still-share-covid-19-hoaxes-157894 [accessed 9 June 2021].

18. World Health Organization. Social Media & COVID-19: A Global Study of Digital Crisis Interaction among Gen Z and Millenials. 6 March 2021. https://www.who.int/news-room/feature-stories/detail/social-media-covid-19-a-global-study-of-digital-crisis-interaction-among-gen-z-and-millennials [accessed 8 August 2020]

19. Tarakini G, Mwedzi T, Manyuchi T, Tarakini T. The role of media during COVID-19 global outbreak: a conservation perspective. Trop Conserv Sci. 2021;14. doi:10.1177/19400829211008088

20. Parvin GA, Ahsan R, Rahman MH, Abedin A. Novel coronavirus (COVID-19) pandemic: the role of printing media in Asian countries. Front Commun. 2020;5(March):1–20. doi:10.3389/fcomm.2020.557593

21. Dhanani LY, Franz B. The role of news consumption and trust in public health leadership in shaping COVID-19 knowledge and prejudice. Front Psychol. 2020;11(October). doi:10.3389/fpsyg.2020.560828

22. Loomba S, de Figueiredo A, Piatek SJ, de Graaf K, Larson HJ. Measuring the impact of COVID-19 vaccine misinformation on vaccination intent in the UK and USA. Nat Hum Behav. 2021;5(3):337–348. doi:10.1038/s41562-021-01056-1

23. Ceniti C, Tilocca B, Britti D, Santoro A, Costanzo N. Food safety concerns in “COVID-19 Era.” Microbiol Res (Pavia). 2021;12(1):53–68. doi:10.3390/microbiolres12010006

24. La Rosa G, Bonadonna L, Lucentini L, Kenmoe S, Suffredini E. Coronavirus in water environments: Occurrence, persistence and concentration methods - A scoping review. Water Res. 2020;179:115899. doi:10.1016/j.watres.2020.115899

25. Morawska L, Milton DK. It is time to address airborne transmission of coronavirus disease 2019 (COVID-19). Clin Infect Dis. 2020;71(9):2311–2313. doi:10.1093/cid/ciaa939

26. The Lancet Respiratory Medicine. COVID-19 transmission—up in the air. Lancet Respir Med. 2020;8(12):1159. doi:10.1016/S2213-2600(20)30514-2

27. Bhanot D, Singh T, Verma SK, Sharad S. Stigma and discrimination during COVID-19 pandemic. Front Public Health. 2021;8:5770188. doi:10.3389/fpubh.2020.577018

28. Turner-Musa J, Ajayi O, Kemp L. Examining social determinants of health, stigma, and COVID-19 disparities. Healthcare (Basel) 2020;8(2):168. doi: 10.3390/healthcare8020168

29. Alzyood M, Jackson D, Aveyard H, Brooke J. COVID-19 reinforces the importance of handwashing. J Clin Nurs. 2020;29(15-16):2760–2761. doi:10.1111/jocn.15313

30. Freeman MC, Caruso BA. Comment on “Global access to handwashing: implications for COVID-19 control in low-income countries”. Environ Health Perspect. 2020;128(9):1–2. doi:10.1289/EHP7852

31. Bikbov B, Bikbov A. Maximum incubation period for COVID-19 infection: do we need to rethink the 14-day quarantine policy? Travel Med Infect Dis. 2020;(January):2020–2022. doi:10.1016/j.tmaid.2021.101976

32. Li ZY, Zhang Y, Peng LQ, Gao RR, Jing JR, Wang JL, et al. Demand for longer quarantine period among common and uncommon COVID-19 infections: A scoping review. Infect Dis Poverty. 2021;10(1):1–9. doi:10.1186/s40249-021-00847-y

33. Jain R, Dupas P. The Effects of India’s COVID-19 Lockdown on Critical Non-COVID Health Care and Outcomes. Working Paper Series on Health and Demographic Change in the Asia-Pacific. 23 September 2020. Asia Health Policy Program working paper #60. Available: https://fsi-live.s3.us-west-1.amazonaws.com/s3fs-public/ahpp_wp_60.pdf [accessed 10 June 2021].

34. Burger R, Day C, Deghaye N, Nkonki L, Rensburg R, Smith A, et al. Examining the Unintended Consequences of the COVID-19 Pandemic on Public Sector Health Facility Visits: The First 150 Days. 2020. Available: https://cramsurvey.org/wp-content/uploads/2020/12/16.-Examining-the-unintended-consequences-of-the-COVID-19-pandemic-on-public-sector-health-facility-visits-The-first-150-days-2.pdf [accessed 10 June 2021].

35. Kanozia R, Arya R. “Fake news”, religion, and COVID-19 vaccine hesitancy in India, Pakistan, and Bangladesh. Media Asia. 2021;0(0):1–9. doi:10.1080/01296612.2021.1921963

36. Schwarzinger M, Luchini S. Addressing COVID-19 vaccine hesitancy: is official communication the key? Lancet Public Heal. 2021;6(6):e353–e354. doi:10.1016/s2468-2667(21)00108-0

37. Freeman D. COVID vaccine hesitancy: spell out the personal rather than collective benefits to persuade people — new research. The Conversation (13 May 2021). Available: https://theconversation.com/covid-vaccine-hesitancy-spell-out-the-personal-rather-than-collective-benefits-to-persuade-people-new-research-160824 [accessed 10 June 2021].

38. Parthasarathi A, Puvvada RK, Shankar M, Siddaiah JB, Ganguly K, Upadhyay S et al. Willingness to accept the COVID-19 vaccine and related factors among Indian adults: a cross-sectional study. Vaccines. 2022;10(7) doi: 10.3390/vaccines10071095

39. Lazarus JV, Wyka K, White TM, Picchio CA, Rabin K, Ratzan SC et al. Revisiting COVID-19 vaccine hesitancy around the world using data from 23 countries in 2021. Nature Commun. 2022:13, 3801. https://doi.org/10.1038/s41467-022-31441-x

40. Dzinamarira T, Nachipo B, Phiri B, Musuka G. COVID-19 vaccine roll-out in South Africa and Zimbabwe: urgent need to address community preparedness, fears and hesitancy. Vaccines. 2021:9(3). https://doi.org/10.3390/vaccines9030250

41. Hoare J, Mendelson M, Frenkel L. COVID-19 vaccine hesitancy and anti-vaxxers – supporting healthcare workers to navigate the unvaccinated: reflections from clinical practice. SAMJ. 2022:112(1). http://dx.doi.org/10.7196/SAMJ.2022.v112i1.16208

